# Comparative Outcomes of Percutaneous Coronary Intervention in Academic Versus Non-Academic Institutions: Insights from a Nationwide Cohort Analysis

**DOI:** 10.1101/2025.10.06.25337450

**Authors:** Muhammad Raffey Shabbir, Malik Ahsan Safdar, Muhammad Osama, Syed Muhammad Salman Hassan, Ahmad Baig, Aneesa Altaf, Khubaib Ahmad, Muhammad Shoaib Khan

**Author notes:** Corresponding author Muhammad Raffey Shabbir, MD | | Marshfield Clinic, Sanford Health, Marshfield, Wisconsin, USA. **Statements and Declarations**. **Ethical Approval:** No ethical approval was required for the study. **Consent:** No consent was needed. **Data Availability Statement:** All data generated or analyzed during this study are included in this article. Further inquiries can be directed to the corresponding author.

## Abstract

**Introduction:** Percutaneous Coronary Intervention (PCI) is a cornerstone in managing coronary artery disease. Outcomes may vary by institutional settings, with academic centers often treating higher-risk cases. This study compares adverse PCI outcomes between academic and non-academic hospitals using real-world data.

**Objectives:** Comparison of adverse outcome of PCI in academic versus non-academic setting using real-world data. We hypothesize that academic centers despite their specialized capabilities, exhibit higher adverse event rates due to the complexity of cases and patient risk profiles.

**Methods:** A retrospective cohort study was conducted using the TriNetX US Collaborative Network. Adults (35–90 years), who underwent PCI between Jan 1, 2010 to Jan 1, 2020 were included. Those with congenital heart disease, rheumatic heart disease, ischemic cardiomyopathy, or prior myocardial infarction (MI) were excluded. Groups were stratified by hospital academic status and balanced using 1:1 propensity score matching. Outcomes were assessed within 90 days post-PCI. Patients with a recorded occurrence of the outcome prior to the index event were excluded from each respective outcome analysis. Kaplan-Meier analysis and log-rank tests were used for statistical comparisons with significance set at p<0.05.

**Results:** Academic centers demonstrated significantly higher hazard ratios (HR) for multiple adverse outcomes. HRs (95% CI): all-cause mortality 1.465 (1.314–1.632), cardiac arrest 11.798 (5.453–25.526), complications in total 3.798 (3.065–4.707), systemic thromboembolism 2.052 (1.412–2.984), cerebrovascular infarction 4.028 (1.137–14.274), acute coronary thrombosis not resulting in MI 2.554 (1.405–4.643), Dressler’s syndrome/pericarditis 2.374 (1.346–4.187), coronary artery aneurysm or dissection 4.560 (3.029–6.866), injury to the radial or femoral artery 27.277 (3.707–200.732), acute kidney injury 3.266 (2.825–3.777), and hematomas/hemorrhages 3.835 (2.864–5.136). MI type 2/4 had a less definitive HR of 1.658 (0.853–3.221).

**Conclusions:** In this retrospective analysis of TriNetX data, for the majority of PCI associated outcomes, statistically significant higher rate of adverse outcomes in academic institution have been observed as compared to non-academic settings. These findings could be due to referral of high risk, and complex cases to academic centers. Future studies should investigate modifiable institutional factors and patient-level variables driving this disparity.

## Introduction

Coronary artery disease is one of the leading causes of mortality and morbidity worldwide. It has been reported that coronary artery disease is the single most important cause of mortality and early life disability in the world. Almost 7 million deaths occur yearly from coronary artery disease in addition to 129 million early life disabilities reported globally (1).

Percutaneous coronary intervention (PCI) is one of the most important and lifesaving procedures. If performed within the given time frame, it can save the lives of patients suffering from acute coronary syndrome. PCI can reduce mortality and recurrence of MI in patients with unstable coronary artery disease. For patients with stable coronary artery disease, PCI shows no evidence of an effect on outcomes provided left coronary artery is involved (2).

Procedural outcomes including mortality, periprocedural complications, and revascularization serve as markers of patient safety and healthcare quality. However, significant disparities exist between different healthcare facilities. Hospital academic status, clinician expertise, available resources, and infrastructure are key contributing factors distinguishing academic from non-academic centers.

While previous studies emphasize the role of PCI in reducing mortality in acute coronary syndrome, little literature exists comparing PCI outcomes in academic versus non-academic hospitals. This gap underscores the need for a nationwide analysis, incorporating real-world data to evaluate outcome disparities. The purpose of this study was to compare PCI outcomes in academic and non-academic hospitals across the United States.

## METHODS

### Data Availability

All data generated or analyzed during this study are included in this article and its supplementary materials. Further inquiries can be directed to the corresponding author.

### Data Source and Study Design

We conducted a retrospective cohort study using the TriNetX, LLC platform, a global federated health research network that provides access to de-identified electronic health records (EHRs) from participating healthcare organizations (HCOs). TriNetX aggregates data across diverse care settings, including hospitals, primary care clinics, and specialty practices, and includes records from both insured and uninsured patients. Available data include demographics, diagnoses (ICD-10-CM), procedures (ICD-10-PCS, CPT), medications (Veterans Affairs National Formulary), laboratory tests (LOINC), genomic data (HGVS), and healthcare utilization metrics.

Our analysis was conducted using the TriNetX US Collaborative Network, which, at the time of data extraction in May 2025, comprised 47 HCOs and 76,822,022 de-identified patients. Natural language processing (NLP) is applied within the platform to enhance structured data fields.

### Ethics Statement

The TriNetX platform complies with the Health Insurance Portability and Accountability Act (HIPAA) and the General Data Protection Regulation (GDPR). Because TriNetX only provides aggregated counts and statistical summaries of de-identified data, individual patient consent is not required. The Western Institutional Review Board (WIRB) has granted a waiver of informed consent for studies utilizing TriNetX data. Ethical approval was deemed unnecessary, consistent with Section §164.514 of the HIPAA Privacy Rule.

### Data Quality

TriNetX ensures data quality through routine evaluations of conformance, completeness, and plausibility, utilizing both internal and external validation methods. These quality assurance processes support its utility in real-world evidence generation and randomized controlled trial (RCT) planning. Additional platform details are provided in the supplementary materials.

### Cohort Definition and Eligibility

Cohort construction followed a two-step process: 1) Defining eligibility criteria using structured query parameters 2) Executing built-in analysis tools within the TriNetX platform. From the US Collaborative Network, utilizing CPT codes (1021163, 1021164, 1021165, 1021166, 1021167, 1021168), we identified adult patients aged 35 to 90 years who underwent percutaneous coronary intervention (PCI) between January 2010 and January 2020 (n = 164,400). We excluded patients with the following ICD-10-CM codes: Chronic rheumatic heart disease (I05–I09), Old myocardial infarction (I25.2), Ischemic cardiomyopathy (I25.5), Other chronic ischemic heart disease (I25.8), and Congenital malformations of the circulatory system (Q20–Q28).

After exclusions (n = 108,729), a final cohort of 55,671 patients was identified and stratified into two groups based on the academic status of the institution where the PCI was performed. Classification of centers as academic or non-academic was determined by TriNetX’s internal metadata. No external or standardized definition was applied: Academic centers: n = 21,331; Non-academic centers: n = 34,340. A cohort construction diagram is provided in Figure 1.

**Figure 1.**
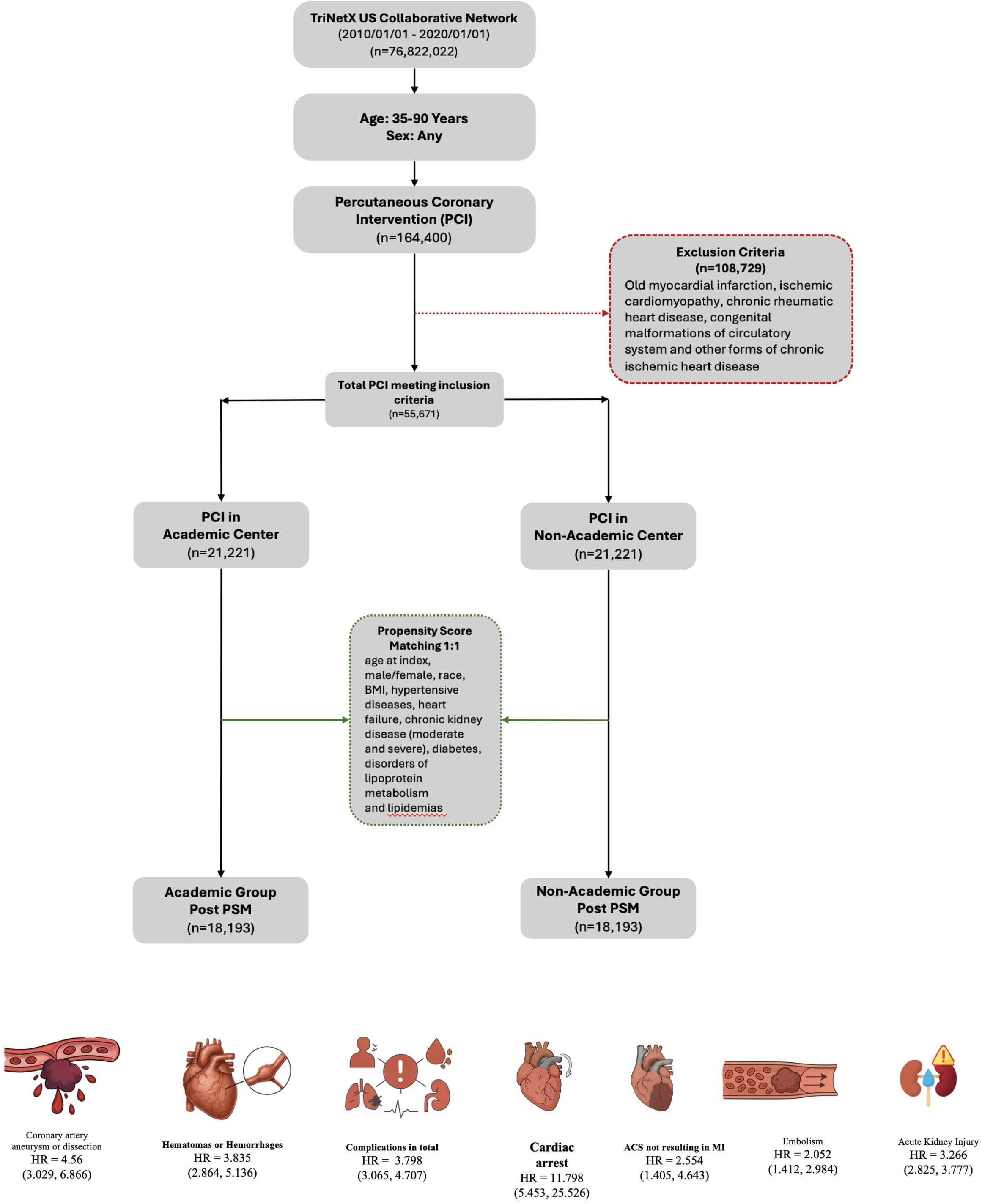
Central Illustration of study design and outcomes.

**Figure 2.**
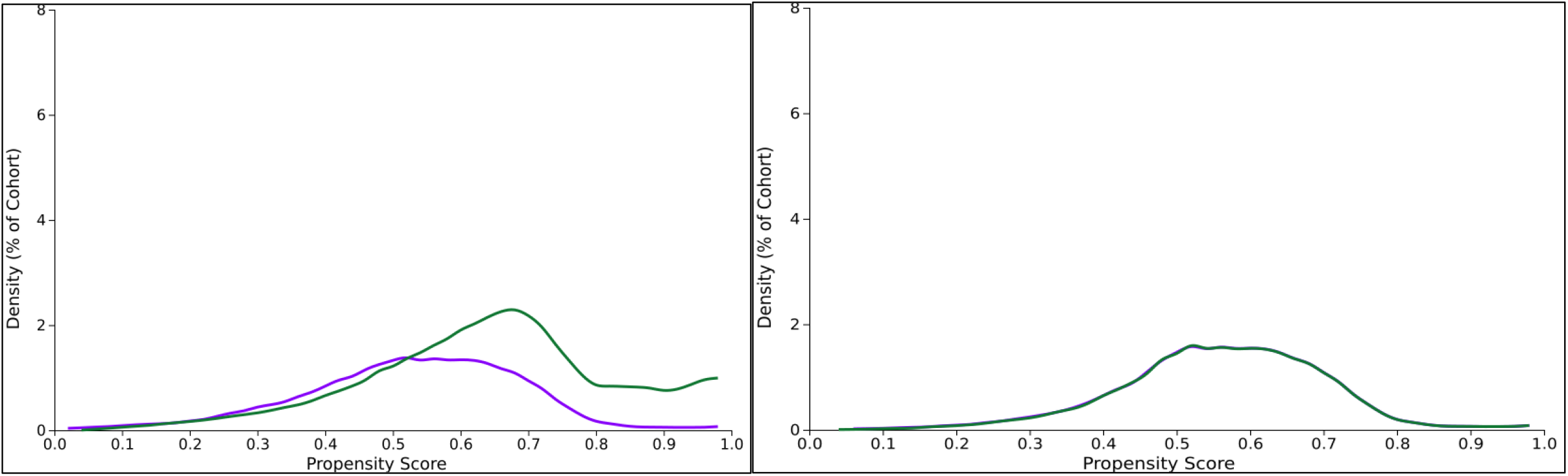
Propensity score density function - Before and after matching (cohort 1 - purple, cohort 2 - green)

**Figure 3.**
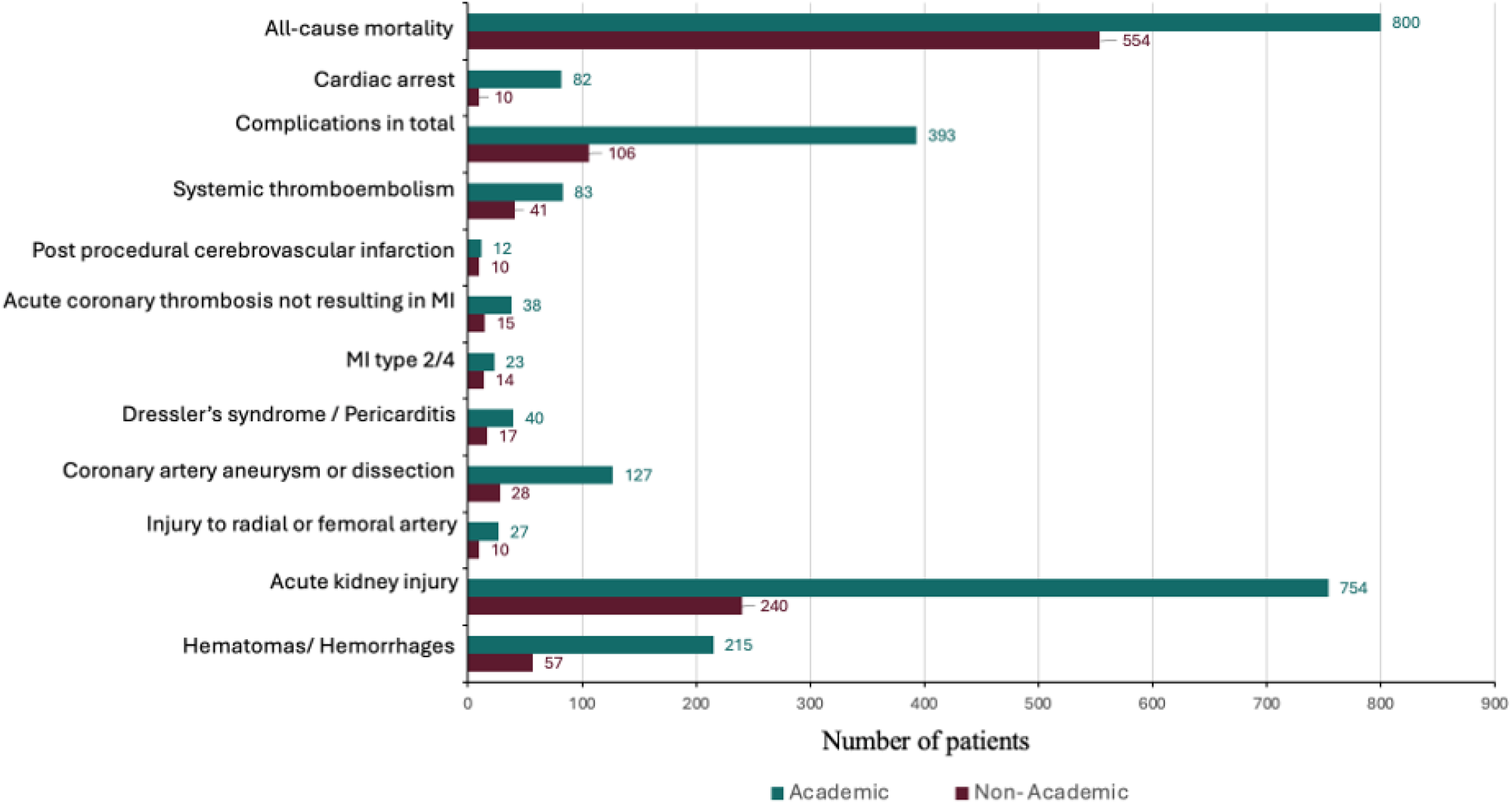
Outcomes in academic versus non-academic institutions after propensity score matching.

**Figure 4.**
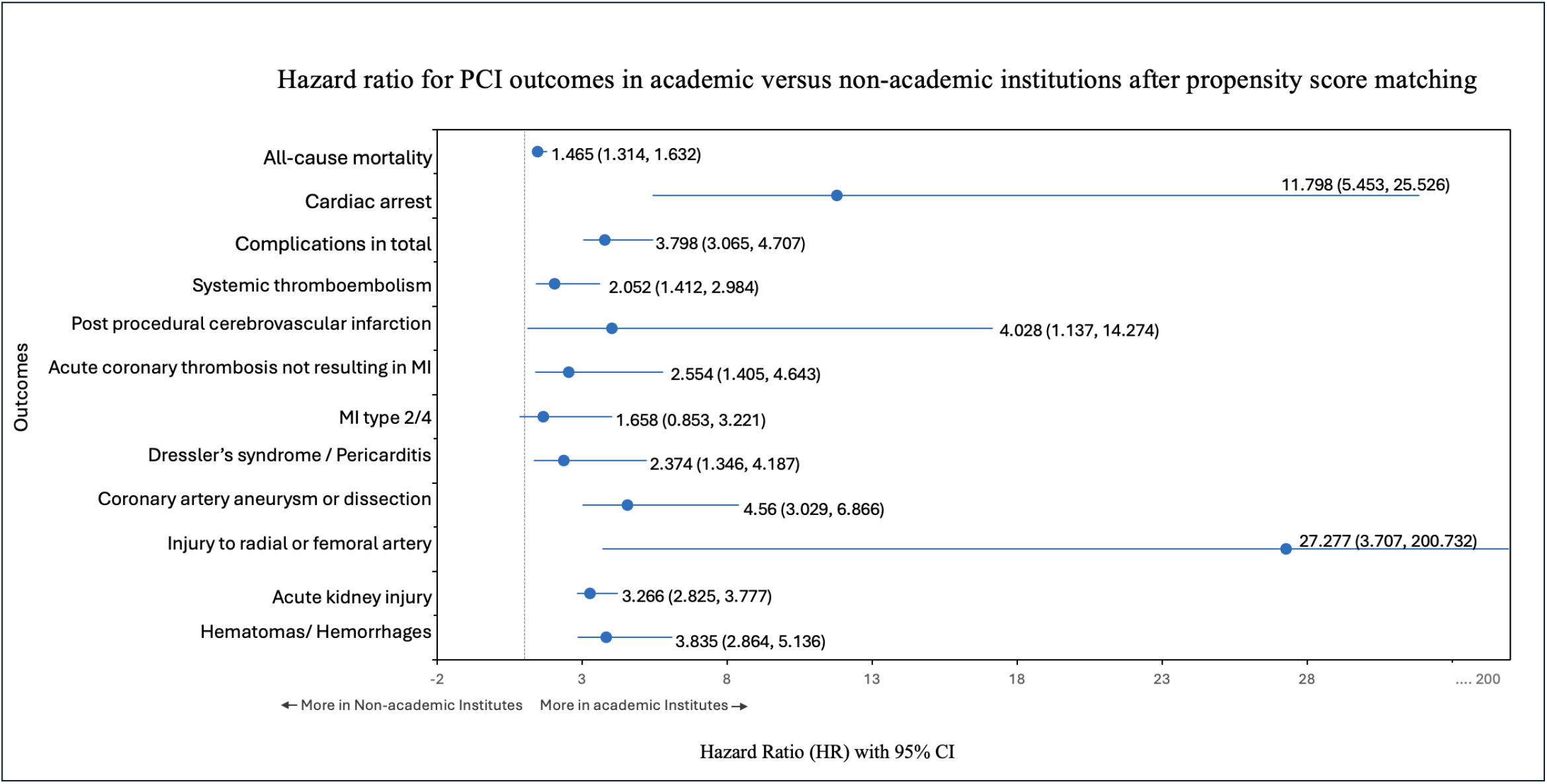
Forest Plot Hazard-Ratio for main cohort analysis.

### Propensity Score Matching (PSM) and Baseline Characteristics

To reduce immortal time bias, the index event was defined as the date on which each patient met all inclusion criteria. To simulate a randomized design and minimize confounding, we applied 1:1 propensity score matching using a greedy nearest neighbor algorithm without replacement and a caliper width of 0.1 based on the pooled standard deviation of the logit of the propensity score. Patients who could not be matched were excluded.

A total of 20 baseline covariates were used for PSM, including: Demographics (age at index, gender, race, ethnicity) and comorbidities (hypertension, atrial fibrillation/flutter, heart failure, disorders of lipoprotein metabolism and lipidemia, diabetes mellitus, chronic kidney disease, and obesity). Covariate balance was evaluated using standardized mean differences (SMD), with an SMD < 0.1 indicating acceptable balance. Density plots and pre-vs. post-match distributions are included in the Supplemental Figures and Tables. Following matching, 18,193 patients remained in each cohort for comparative outcome analysis.

### Outcomes and Endpoints

The study outcomes were assessed during a 90-day follow-up period following the index PCI. Outcomes were identified using validated CPT and ICD-10 codes, including: all-cause mortality, intraoperative cardiac arrest (I97.71), overall postprocedural complications (I97.x), systemic thromboembolism (I74), postprocedural cerebrovascular infarction (I97.82), acute coronary thrombosis not resulting in myocardial infarction (I24), myocardial infarction – Type 2 or Type 4 (I21.A1), Dressler’s Syndrome/acute pericarditis (I24.1, I30), coronary artery aneurysm or dissection (I25.4), injury to radial or femoral arteries (S65.1, S75.x), acute kidney injury (N17.x), and postprocedural/intraprocedural hematomas or hemorrhages (I97.4x, I97.5x, I97.6x).

### Statistical Analysis

All statistical analyses were performed using the built-in analytic tools of the TriNetX platform. Continuous variables were presented as means ± standard deviations and compared using independent samples t-tests, while categorical variables were summarized as counts and percentages and compared using Chi-square tests.

Survival outcomes were analyzed using the Kaplan-Meier method, and survival distributions between groups were compared using the log-rank test. Hazard ratios (HRs) with 95% confidence intervals (CIs) were calculated to assess the relative risk of outcomes in patients undergoing PCI at academic versus non-academic institutions. The log-rank test statistic was interpreted using a chi-square distribution with 1 degree of freedom (df = 1), as appropriate for two-group comparisons.

To estimate measures of association between the matched cohorts, we used risk ratios (RRs), odds ratios (ORs), and absolute risk differences (RDs), each reported with corresponding 95% CIs. These were calculated directly within the platform’s outcome comparison tools (see Supplementary Table). A two-sided p-value < 0.05 was considered statistically significant. All analyses were conducted entirely within the TriNetX platform; no data were exported for external statistical analysis.

### Sensitivity Analyses

Three sensitivity analyses were conducted to assess the robustness of the study findings:

1. Exclusion of Baseline High-Risk Patients: The first analysis re-evaluated the matched cohorts after excluding individuals with pre-existing conditions known to influence PCI outcomes, such as severe valvular disease, advanced malignancy, or coagulopathy.
2. Pre-vs. Post-PSM Comparison: The second analysis assessed the consistency of effect estimates before and after propensity score matching by applying the same analytic approach to the unmatched (pre-PSM) population. This validated that the matching procedure did not distort the direction or significance of observed outcomes.

Further methodological specifications, including variable definitions and diagnostics for covariate balance, are available in tables 1-3.

**Table 1:**
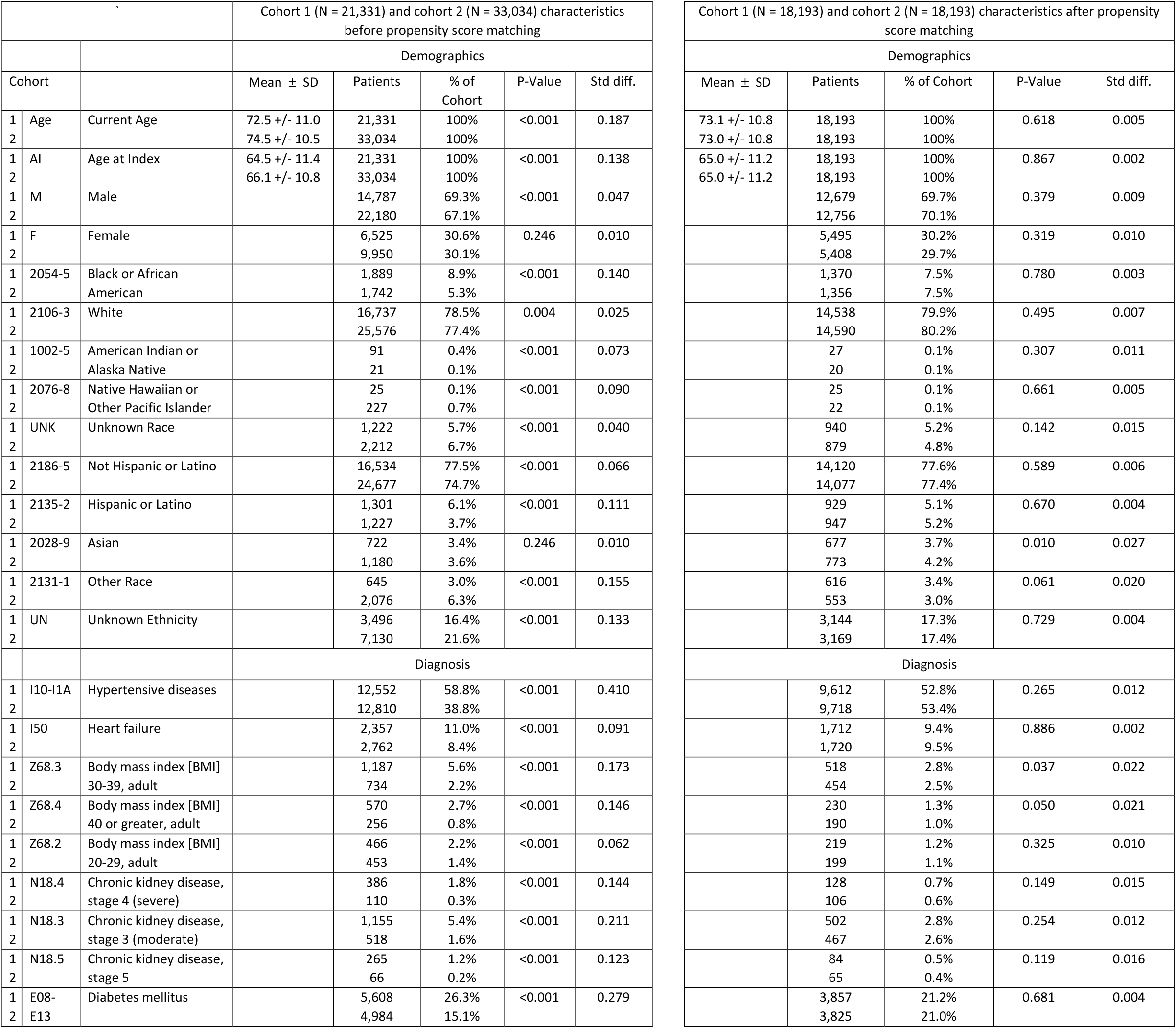
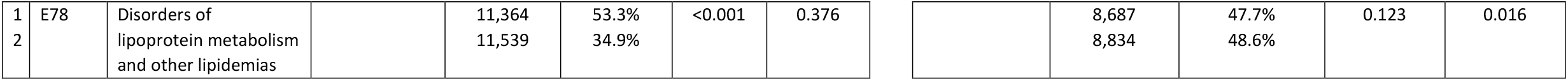
Baseline Characteristics.

|  |  |  | Cohort 1 (N = 21,331) and cohort 2 (N = 33,034) characteristics before propensity score matching |  |  |  |  |
| --- | --- | --- | --- | --- | --- | --- | --- |
|  |  |  | Demographics |  |  |  |  |
| Cohort |  |  | Mean ± SD | Patients | % of Cohort | P-Value | Std diff. |
| 1 | Age | Current Age | 72.5 +/- 11.0 | 21,331 | 100% | <0.001 | 0.187 |
| 2 |  |  | 74.5 +/- 10.5 | 33,034 | 100% |  |  |
| 1 | AI | Age at Index | 64.5 +/- 11.4 | 21,331 | 100% | <0.001 | 0.138 |
| 2 |  |  | 66.1 +/- 10.8 | 33,034 | 100% |  |  |
| 1 | M | Male |  | 14,787 | 69.3% | <0.001 | 0.047 |
| 2 |  |  |  | 22,180 | 67.1% |  |  |
| 1 | F | Female |  | 6,525 | 30.6% | 0.246 | 0.010 |
| 2 |  |  |  | 9,950 | 30.1% |  |  |
| 1 | 2054-5 | Black or African American |  | 1,889 | 8.9% | <0.001 | 0.140 |
| 2 |  |  |  | 1,742 | 5.3% |  |  |
| 1 | 2106-3 | White |  | 16,737 | 78.5% | 0.004 | 0.025 |
| 2 |  |  |  | 25,576 | 77.4% |  |  |
| 1 | 1002-5 | American Indian or Alaska Native |  | 91 | 0.4% | <0.001 | 0.073 |
| 2 |  |  |  | 21 | 0.1% |  |  |
| 1 | 2076-8 | Native Hawaiian or Other Pacific Islander |  | 25 | 0.1% | <0.001 | 0.090 |
| 2 |  |  |  | 227 | 0.7% |  |  |
| 1 | UNK | Unknown Race |  | 1,222 | 5.7% | <0.001 | 0.040 |
| 2 |  |  |  | 2,212 | 6.7% |  |  |
| 1 | 2186-5 | Not Hispanic or Latino |  | 16,534 | 77.5% | <0.001 | 0.066 |
| 2 |  |  |  | 24,677 | 74.7% |  |  |
| 1 | 2135-2 | Hispanic or Latino |  | 1,301 | 6.1% | <0.001 | 0.111 |
| 2 |  |  |  | 1,227 | 3.7% |  |  |
| 1 | 2028-9 | Asian |  | 722 | 3.4% | 0.246 | 0.010 |
| 2 |  |  |  | 1,180 | 3.6% |  |  |
| 1 | 2131-1 | Other Race |  | 645 | 3.0% | <0.001 | 0.155 |
| 2 |  |  |  | 2,076 | 6.3% |  |  |
| 1 | UN | Unknown Ethnicity |  | 3,496 | 16.4% | <0.001 | 0.133 |
| 2 |  |  |  | 7,130 | 21.6% |  |  |
|  |  |  | Diagnosis |  |  |  |  |
| 1 | I10-I1A | Hypertensive diseases |  | 12,552 | 58.8% | <0.001 | 0.410 |
| 2 |  |  |  | 12,810 | 38.8% |  |  |
| 1 | I50 | Heart failure |  | 2,357 | 11.0% | <0.001 | 0.091 |
| 2 |  |  |  | 2,762 | 8.4% |  |  |
| 1 | Z68.3 | Body mass index [BMI] 30-39, adult |  | 1,187 | 5.6% | <0.001 | 0.173 |
| 2 |  |  |  | 734 | 2.2% |  |  |
| 1 | Z68.4 | Body mass index [BMI] 40 or greater, adult |  | 570 | 2.7% | <0.001 | 0.146 |
| 2 |  |  |  | 256 | 0.8% |  |  |
| 1 | Z68.2 | Body mass index [BMI] 20-29, adult |  | 466 | 2.2% | <0.001 | 0.062 |
| 2 |  |  |  | 453 | 1.4% |  |  |
| 1 | N18.4 | Chronic kidney disease, stage 4 (severe) |  | 386 | 1.8% | <0.001 | 0.144 |
| 2 |  |  |  | 110 | 0.3% |  |  |
| 1 | N18.3 | Chronic kidney disease, stage 3 (moderate) |  | 1,155 | 5.4% | <0.001 | 0.211 |
| 2 |  |  |  | 518 | 1.6% |  |  |
| 1 | N18.5 | Chronic kidney disease, stage 5 |  | 265 | 1.2% | <0.001 | 0.123 |
| 2 |  |  |  | 66 | 0.2% |  |  |
| 1 | E08-E13 | Diabetes mellitus |  | 5,608 | 26.3% | <0.001 | 0.279 |
| 2 |  |  |  | 4,984 | 15.1% |  |  |

| Cohort 1 (N = 18,193) and cohort 2 (N = 18,193) characteristics after propensity score matching |  |  |  |  |
| --- | --- | --- | --- | --- |
| Demographics |  |  |  |  |
| Mean ± SD | Patients | % of Cohort | P-Value | Std diff. |
| 73.1 +/- 10.8 | 18,193 | 100% | 0.618 | 0.005 |
| 73.0 +/- 10.8 | 18,193 | 100% |  |  |
| 65.0 +/- 11.2 | 18,193 | 100% | 0.867 | 0.002 |
| 65.0 +/- 11.2 | 18,193 | 100% |  |  |
|  | 12,679 | 69.7% | 0.379 | 0.009 |
|  | 12,756 | 70.1% |  |  |
|  | 5,495 | 30.2% | 0.319 | 0.010 |
|  | 5,408 | 29.7% |  |  |
|  | 1,370 | 7.5% | 0.780 | 0.003 |
|  | 1,356 | 7.5% |  |  |
|  | 14,538 | 79.9% | 0.495 | 0.007 |
|  | 14,590 | 80.2% |  |  |
|  | 27 | 0.1% | 0.307 | 0.011 |
|  | 20 | 0.1% |  |  |
|  | 25 | 0.1% | 0.661 | 0.005 |
|  | 22 | 0.1% |  |  |
|  | 940 | 5.2% | 0.142 | 0.015 |
|  | 879 | 4.8% |  |  |
|  | 14,120 | 77.6% | 0.589 | 0.006 |
|  | 14,077 | 77.4% |  |  |
|  | 929 | 5.1% | 0.670 | 0.004 |
|  | 947 | 5.2% |  |  |
|  | 677 | 3.7% | 0.010 | 0.027 |
|  | 773 | 4.2% |  |  |
|  | 616 | 3.4% | 0.061 | 0.020 |
|  | 553 | 3.0% |  |  |
|  | 3,144 | 17.3% | 0.729 | 0.004 |
|  | 3,169 | 17.4% |  |  |
| Diagnosis |  |  |  |  |
|  | 9,612 | 52.8% | 0.265 | 0.012 |
|  | 9,718 | 53.4% |  |  |
|  | 1,712 | 9.4% | 0.886 | 0.002 |
|  | 1,720 | 9.5% |  |  |
|  | 518 | 2.8% | 0.037 | 0.022 |
|  | 454 | 2.5% |  |  |
|  | 230 | 1.3% | 0.050 | 0.021 |
|  | 190 | 1.0% |  |  |
|  | 219 | 1.2% | 0.325 | 0.010 |
|  | 199 | 1.1% |  |  |
|  | 128 | 0.7% | 0.149 | 0.015 |
|  | 106 | 0.6% |  |  |
|  | 502 | 2.8% | 0.254 | 0.012 |
|  | 467 | 2.6% |  |  |
|  | 84 | 0.5% | 0.119 | 0.016 |
|  | 65 | 0.4% |  |  |
|  | 3,857 | 21.2% | 0.681 | 0.004 |
|  | 3,825 | 21.0% |  |  |

|  |  |  |  |  |  |  |  |  |  |  |  |  |
| --- | --- | --- | --- | --- | --- | --- | --- | --- | --- | --- | --- | --- |
| 1 | E78 | Disorders of lipoprotein metabolism and other lipidemias |  | 11,364 | 53.3% | <0.001 | 0.376 |  | 8,687 | 47.7% | 0.123 | 0.016 |
| 2 |  |  |  | 11,539 | 34.9% |  |  |  | 8,834 | 48.6% |  |  |

**Table 4:** Propensity score matching.

| Cohort | Patient count before matching | Patient count after matching |
| --- | --- | --- |
| 1 - Academic | 21,331 | 18,193 |
| 2 - Non-Academic | 33,034 | 18,193 |

**Table 5:** Hazard ratio, risk ratio, and risk difference for PCI outcomes in academic versus non-academic institutions after propensity score matching.

| Outcomes | Hazard Ratio<br>HR (95% CI) | Kaplan-Meier<br>log-rank test |  |  | Risk ratio<br>RR (95% CI) | Odds Ratio<br>OR (95% CI) | Risk Difference<br>RD (95% CI) |
| --- | --- | --- | --- | --- | --- | --- | --- |
|  |  | χ<sup>2</sup> | df | p-value |  |  |  |
| All-cause mortality | 1.465 (1.314, 1.632) | 48.257 | 1 | <0.001 | 1.445 (1.299, 1.607) | 1.463 (1.310, 1.633) | 0.012 (0.008, 0.015) |
| Cardiac arrest | 11.798 (5.453, 25.526) | 63.77 | 1 | <0.001 | 8.215 (4.262, 15.835) | 8.244 (4.275, 15.899) | 0.003 (0.003, 0.004) |
| Complications in total | 3.798 (3.065, 4.707) | 172.318 | 1 | <0.001 | 3.75 (3.029, 4.643) | 3.804 (3.067, 4.719) | 0.014 (0.012, 0.016) |
| Systemic thromboembolism | 2.052 (1.412, 2.984) | 14.811 | 1 | <0.001 | 2.033 (1.399, 2.954) | 2.037 (1.400, 2.963) | 0.002 (0.001, 0.003) |
| Post procedural cerebrovascular infarction | 4.028 (1.137, 14.274) | 5.464 | 1 | 0.019 | 1.2 (0.519, 2.777) | 1.2 (0.518, 2.779) | 0 (-0.000, 0.001) |
| Acute coronary thrombosis not resulting in MI | 2.554 (1.405, 4.643) | 10.169 | 1 | 0.001 | 2.536 (1.395, 4.608) | 2.539 (1.396, 4.616) | 0.001 (0.000, 0.002) |
| MI type 2/4 | 1.658 (0.853, 3.221) | 2.27 | 1 | 0.132 | 1.645 (-0.000, 0.001) | 1.645 (0.847, 3.199) | 0 (-0.000, 0.001) |
| Dressler’s syndrome / Pericarditis | 2.374 (1.346, 4.187) | 9.487 | 1 | 0.002 | 2.354 (1.335, 4.151) | 2.357 (1.336, 4.158) | 0.001 (0.000, 0.002) |
| Coronary artery aneurysm or dissection | 4.56 (3.029, 6.866) | 63.846 | 1 | <0.001 | 4.543 (3.019, 6.837) | 4.565 (3.031, 6.876) | 0.005 (0.004, 0.006) |
| Injury to radial or femoral artery | 27.277 (3.707, 200.732) | 24.41 | 1 | <0.001 | 2.701 (1.308, 5.578) | 2.703 (1.308, 5.585) | 0.001 (0.000, 0.001) |
| Acute kidney injury | 3.266 (2.825, 3.777) | 287.347 | 1 | <0.001 | 3.217 (2.786, 3.716) | 3.306 (2.855, 3.828) | 0.026 (0.023, 0.029) |
| Hematomas/ Hemorrhages | 3.835 (2.864, 5.136) | 94.49 | 1 | <0.001 | 3.796 (2.837, 5.080) | 3.826 (2.855, 5.126) | 0.008 (0.006, 0.009) |

Abbreviations/Key: CI = Confidence interval | df = Degrees of freedom | HR = Hazard ratio | RD = Risk difference | RR = Risk ratio | OR = Odds ratio | PCI = Percutaneous Coronary Intervention | MI = Myocardial infarction

## Results

This study aimed to determine whether the type of institution, academic versus non-academic affects clinical outcomes in patients undergoing percutaneous coronary intervention (PCI).

Before propensity score matching, the two cohorts were imbalanced. The academic cohort initially included 21,331 patients, while the non-academic cohort had 33,034 patients. The non-academic group was older on average, with a mean current age of 74.5 ± 10.5 years, compared to the academic group’s mean age of 72.5 ± 11.0 years (P < 0.001). There were also notable differences in race and ethnicity. The academic cohort had 1,889 Black or African American patients (8.9%) versus 1,742 (5.3%) in the non-academic group (P < 0.001).

The academic cohort also showed a higher prevalence of hypertensive diseases (58.8% vs. 38.8%) (P < 0.001), heart failure (11.0% vs. 8.4%) (P < 0.001), diabetes mellitus (40.1% vs. 36.0%) (P < 0.001), myocardial infarction (7.1% vs. 6.5%) (P=0.038), chronic kidney disease (21.8% vs. 20.4%) (P < 0.001), and cardiomyopathy (1.3% vs. 1.0%) (P < 0.001).

To address these baseline differences, propensity score matching was used to create two balanced cohorts, each with 18,193 patients. After matching, the mean current age was 73.1 ± 10.8 years in the academic group and 73.0 ± 10.8 years in the non-academic group (P=0.618). Similarly, the prevalence of conditions like hypertensive diseases (52.8% in academic vs. 53.4% in non-academic; P=0.265) and heart failure (9.4% in academic vs. 9.5% in non-academic; P=0.886) were no longer statistically different between the two groups.

The analysis of the matched cohorts indicated a statistically significant increased risk for most adverse outcomes at academic institutions. All-cause mortality occurred in 800 patients (4.4%) in the academic group and 554 (3.0%) in the non-academic group, with a Hazard Ratio (HR) of 1.465 (95% CI: 1.314, 1.632; P < 0.001), a Risk Ratio (RR) of 1.445 (95% CI: 1.299, 1.607), and an Odds Ratio (OR) of 1.463 (95% CI: 1.310, 1.633). For In-Hospital Mortality, there were 434 academic events (2.4%) versus 288 non-academic events (1.6%), with an RR of 1.507 (95% CI: 1.306, 1.738; P < 0.001). Cardiac arrest occurred in 82 academic patients (0.5%) and 10 non-academic patients (0.1%), resulting in an HR of 11.798 (95% CI: 5.453, 25.526; P < 0.001), an RR of 8.215 (95% CI: 4.262, 15.835), and an OR of 8.244 (95% CI: 4.275, 15.899). Total complications were seen in 393 academic patients (2.2%) versus 106 (0.6%) non-academic patients, with an HR of 3.798 (95% CI: 3.065, 4.707; P < 0.001), an RR of 3.75 (95% CI: 3.029, 4.643), and an OR of 3.804 (95% CI: 3.067, 4.719). For Acute Kidney Injury, there were 754 events (4.1%) at academic institutions and 240 (1.3%) at non-academic institutions, with an HR of 3.266 (95% CI: 2.825, 3.777; P < 0.001), an RR of 3.217 (95% CI: 2.786, 3.716), and an OR of 3.306 (95% CI: 2.855, 3.828). Hematomas/Hemorrhages occurred in 215 academic patients (1.2%) versus 57 (0.3%) non-academic patients, with an HR of 3.835 (95% CI: 2.864, 5.136; P < 0.001), an RR of 3.796 (95% CI: 2.837, 5.080), and an OR of 3.826 (95% CI: 2.855, 5.126). Acute Coronary Thrombosis was seen in 38 academic patients (0.2%) and 15 non-academic patients (0.1%), with an HR of 2.554 (95% CI: 1.405, 4.643; P=0.001). Post Procedural Cerebrovascular Infarction occurred in 12 academic patients (0.1%) and 10 non-academic patients (0.1%), with an HR of 4.028 (95% CI: 1.137, 14.274; P=0.019).

In contrast, a few outcomes did not show a statistically significant difference. MI Type 2/4 occurred in 23 academic patients (0.1%) and 14 non-academic patients (0.1%), but its HR of 1.658 (95% CI: 0.853, 3.221) and RR of 1.645 (95% CI: 0.000, 0.001) were not significant, as the confidence interval for the HR crossed 1.0 (P=0.132).

## Discussion

By using the TriNetX database, we found that adult patients undergoing PCI at academic institutions from 2010 to 2020 had a statistically significant hazard ratio of having various complications compared to non-academic centres. The complications ranged from common, like all-cause mortality, cardiac arrest and systemic thromboembolism, to rare conditions like Dressler’s Syndrome and carotid artery dissection/aneurysm. These results are unexpected and opposite to the common belief that academic institutions provide better outcomes.

One of the reasons for these results is that many complex and high-risk patients may get more commonly referred to academic institutions as compared to non-academic centres. Previous studies have shown that teaching hospitals perform more complex PCIs, such as those involving cardiogenic shock, left main disease, or chronic total occlusions [3]. We did a propensity score matching to match baselines characteristics as much as possible but certain things like complexity of case, severity of comorbidities and urgency in procedure cannot be accounted for in datasets.

Another possible explanation can be the changes in personnel in academic institutions. Every year in July, academic institutions get new fellows who are then trained into future cardiologists. But at the start of their training, their involvement can lead to increased procedural duration and hence increased chances of future complications. Young et al. examined the results of 39 studies assessing the effect of the end-of-year changeover on quality of care. A trend toward worse mortality and efficiency of care at the time of academic year-end changeovers was seen [4].

Another important aspect to keep in mind is that new techniques and technologies are first incorporated in academic institutions. Even though innovation paves the way for improvement in healthcare, at first there is a learning curve associated with new devices and techniques leading to more complications. Studies have shown that when new PCI tools are implemented, success rates increased significantly as institutions gained experience. Teaching hospitals exhibited slower early success partly due to trainee involvement and large teams [5]. This balance between innovation and complications is very delicate and any disturbance may lead to increased complications.

It’s also possible that academic institutions may be more rigorous in their reporting of complications due to institutional review boards. The difference of workflow structure in academic and non-academic institutions is also noteworthy. In academic centres, there is a complex team hierarchy, and the decision-making pathways are long. This is done to improve safety and oversight, but it can sometimes lead to delays. Whereas non-academic hospitals may benefit from direct communication and direct execution of PCI. [6]

Interestingly, our results are in contrast with previous studies which show comparable or in some cases even better outcomes for PCI in academic centres compared to non-academic institutions. Jenna et all. showed that high risk acute MI patients may have comparable mortality rates only in July, but the rate is lower in academic institutes throughout the rest of year [7]. In a National Inpatient Sample analysis, Chang et all. reported that among urban hospitals, non-teaching hospitals are associated with higher rates of mortality after PCI [8]. Most of these studies are more than 10 years old and that may be the reason for the contrasting results. As with time, not only electronic medical records (EMRs) have improved but also documentation is more detailed. Another explanation for the contrasting results is difference in patient populations and the scope of the dataset.

The strength of our study includes using a vast dataset i.e. TriNetX which takes patient-level data from different healthcare practices offering a more real-world clinical practice look at PCI outcomes. The use of propensity score matching helped minimize selection bias and allowed us to make meaningful comparisons between the two groups.

That said, this study has limitations. We were limited by what TriNetX has, lacking data on lesion anatomy, operator volume or experience, and institutional protocols, all of which can influence outcomes. Additionally, long-term follow-up was not available, so we were unable to assess outcomes beyond the immediate or early post-procedural period.

## Conclusion

Our results show that, in real-world practice, PCI at academic institutions is associated with higher mortality and complication rates than in non-academic settings. This doesn’t mean academic hospitals should be avoided but it does suggest that complexity of care, procedural dynamics, and institutional structures may influence outcomes more than academic status alone. Future research should explore these factors more in-depth so that healthcare delivery can be improved regardless of the hospital setting.

## Data Availability

All data produced in the present work are contained in the manuscript

https://live.trinetx.com/

## Acknowledgments

None

